# The Norwegian Sports Membership Database: coverage, biases and research potential

**DOI:** 10.1101/2025.10.03.25337227

**Authors:** Fredrik Methi, Ingeborg Hess Elgersma, Jonas Minet Kinge, Arnstein Mykletun, Rannveig Kaldager Hart, Atle Fretheim, Karin Magnusson

**Author notes:** **Corresponding author:** Fredrik Methi, Norwegian Institute of Public Health, Department for Health Services, Cluster for Health Services Research, Oslo, Norway. The main objective of this paper is to describe the content of the NSMD, assess its coverage and develop definitions of active memberships.

## Abstract

**Aims:** To describe the Norwegian Sports Membership Database (NSMD), assess its coverage and develop definitions of active sports memberships.

**Methods:** We validated the coverage of NSMD, a database capturing individual-level sports memberships among persons 10 to 70 years of age between 2015 and 2024. We compared yearly membership counts in NSMD with figures from official reports from The Norwegian Olympic and Paralympic Committee and Confederation of Sports (NIF). We defined all memberships as memberships lasting one day or longer, and active memberships as memberships lasting at least one season or with a paid licence.

**Result:** Between 2015 and 2024, NSMD contained 3 148 213 memberships in which 2 180 621 (60%) was considered active. In total, there were 1 606 048 unique individuals across 55 sports federations and 279 disciplines. Football accounted for 24% of all memberships, handball for 11%, and gymnastics for 9%. Database coverage was low in 2015, capturing 11–16% of expected memberships, but improved substantially over time and exceeded 85% of all memberships and 92% of active memberships in 2023 for ages 13–19.

**Conclusion:** The coverage of the NSMD is high in recent years, representing a level of completeness that allows meaningful population-level analyses from 2019, although with large variations between sports. Linking sports data with health and socio-demographic outcomes provides a potentially large opportunity for population-based research on sports participation and health across the lifespan.

## Introduction

Population-based registries, such as health care registries, have been an invaluable resource for researchers, policymakers, and other stakeholders ^1^. They have provided unique opportunities to study large populations over time ^2–4^, to identify risk factors ^5^, and to evaluate interventions in real-world settings ^6^. Such registries have been particularly powerful when they can be linked with other administrative data sources, such as education or tax records, enabling analyses that cut across different domains of society ^7^. However, despite the well-documented value of registries in other fields, no comparable population-level data sources currently exist for organized sports participation. This represents a significant gap, given the potential of such data to inform research on physical activity, social inequality, public health, and policy development. Establishing registries for organized sports could therefore open new avenues for interdisciplinary research and provide an important complement to existing health and administrative data sources.

In this paper we present the Norwegian Sports Membership Database (NSMD) administered by the Norwegian Olympic and Paralympic Committee and Confederation of Sports (NIF). This database covers all recorded memberships in organized sports in Norway between 2015 and 2024. The database provides a unique opportunity to track sports participation over time, providing detailed insights into new, ongoing, and discontinued memberships in more than 50 sports and 200 disciplines. By linking this database to other national registries, researchers can conduct longitudinal studies on sport memberships together with socio-economy and health.

## Methods

### Data source and collection

The NSMD covers all identifiable membership records in organized sports in Norway. Each membership record is linked to an individual’s national identity number. This identity number is unique to all Norwegian residents and is key in allowing integration with other data sources and registries ^8^. NSMD was gradually established from 2015, following NIF’s mandate requiring all sport federations in Norway to adopt membership systems compatible with this centralized database ^9^.

The sample from NSMD used in this description included all sport memberships for members aged 10 to 70 years between January 1st, 2015 and October 31st, 2024. Each record contained detailed information on start date, end date, sport, and discipline. The database contained no missing values but end dates was absent if the membership was still active at the time of data extraction, October 31st, 2024.

For 34 of the 55 sports, information on paid licences were available (S-Table 1). Licences cover insurance and are usually required from the year a child turns 13 in order to play games and compete in tournaments ^10^. The licence data contained each member’s national identity number, sport, and the licence’s expiration date. The start date was defined as one year prior to the expiration date.

To provide a thorough description of the available data, we linked each person to their registered sex, age, county of residence, and country of birth from The Norwegian Population Register ^8^ along with their parents’ income from Statistics Norway ^11^.

### Statistical analysis

We first described all memberships in NSMD by sex, age, country of birth, participation length, and type of sport for individuals aged 10–70 between 2015 and 2024. A membership was defined as anyone registered in at least one organized sport for at least one day during a calendar year in which they were aged 10–70. Memberships were classified as all memberships (lasting *≥* 1 day) or active memberships. Active memberships followed NIF’s definition of competitive or full-season participation ^12^ and were operationalised as memberships with a paid licence or lasting an entire season. We defined seasons in each sport based on their main competitive season: February-November for sports with summer seasons (e.g., football, athletics, cycling, martial arts) and September-June for sports with winter seasons (e.g., handball, skiing, gymnastics, and motor sports). Supplementary Information 1 and S-Table 1 contain a more detailed description of all sports, their seasons, and availability of licenses.

We then assessed the coverage of NSMD by comparing the number of all memberships and the number of active memberships each year with NIF’s annual reports ^12^. NIF reported both a number for all memberships and a number for active memberships. We calculated all memberships on December 31st, each year between 2015 and 2023. For active members, we stratified the coverage by each sports federation. The discrepancy between the numbers found in NSMD and in NIF’s reports comes from how the data are collected. NSMD is based on individual membership records linked to national identity numbers, while NIF’s reports rely on aggregated membership counts reported by federations and clubs.

Because NIF reported totals by age groups and our data covered ages 10–70, we restricted the coverage analysis to individuals aged 13–70. For sport-specific coverage, we focused on ages 13–19, since older age groups often include coaches and administrative members rather than active athletes. We removed company sports, university sports, and multisports (see Supplementary Information 1).

After estimating coverage, we examined whether registered members in low-coverage years differed from those in high-coverage years. Based on already calculated coverage rates, we classified sports and years into four groups: low (0-39%), fair (40-59%), moderate (60-79%), and high (80-100%) coverage, using terminology from Norwegian medical quality registers ^13^. We then investigated variations in age, geography, and socio-economic factors over time within sports, particularly focusing on the youngest age group, 13–19 years, to determine if using data from years with low coverage would induce biases.

All data management was conducted in R version 4.4.1 using Rstudio. R-scripts are available on GitHub together with an example dataset showing the structure of the data files: https://github.com/MethiF/2025_The_Norwegian_Sports_Membership_Database.

### Ethical considerations

Ethical approval for the NorSport-project ^14^ was granted by the Regional Committees for Medical Research Ethics South East Norway on August 23rd, 2023 (reference #584239).

## Results

### Descriptive results

Between 2015 and 2024, there were 3 148 213 registered sports memberships in NSMD (Table 1). Of these, 2 180 621 (60%) were considered active memberships, lasting more than one season or with a paid licence. Males were overrepresented both in terms of all memberships (56%) and active memberships (57%), and most members were born in Norway (92%).

**Table 1.** Descriptive statistics of memberships in NSMD.

| Characteristics | All memberships | Active memberships |
| --- | --- | --- |
| **Total memberships, n (%)** | 3 148 213 (100.0) | 2 180 621 (100.0) |
| **Unique individuals, n (%)** | 1 606 048 (51.0) | 1 301 669 (59.7) |
| **Female, n (%)** | 1 386 822 (44.1) | 932 830 (42.8) |
| **Age categories, n (%)** |  |  |
| Children (10–12 years) | 1 328 433 (42.2) | 886 072 (40.6) |
| Adolescents (13–19 years) | 535 903 (17.0) | 389 238 (17.8) |
| Young adults (20–39 years) | 649 744 (20.6) | 437 486 (20.1) |
| Middle-aged (40–59 years) | 522 529 (16.6) | 384 762 (17.6) |
| Older adults (60–70 years) | 111 604 (3.5) | 83 063 (3.8) |
| **Country of birth, n (%)** |  |  |
| Norway | 2 891 453 (91.8) | 2 008 712 (92.1) |
| **Membership duration** |  |  |
| Mean days (SD) | 947 (855) | 1 271 (831) |
| Median days (IQR) | 675 (296–1 372) | 1 035 (648–1 739) |
| **Duration categories, n (%)** |  |  |
| Very short (1–30 days) | 134 748 (4.3) | 6 998 (0.3) |
| Short (31–90 days) | 184 372 (5.9) | 15 140 (0.7) |
| Medium (91–365 days) | 660 618 (21.0) | 137 412 (6.3) |
| Long (>365 days) | 2 168 475 (68.9) | 2 021 071 (92.7) |
| **Sports, n (%)** |  |  |
| Football | 756 879 (24.0) | 551 034 (25.3) |
| Handball | 343 283 (10.9) | 247 176 (11.3) |
| Gymnastics | 289 525 (9.2) | 145 599 (6.7) |
| Skiing | 218 651 (6.9) | 156 539 (7.2) |
| Athletics | 138 905 (4.4) | 100 895 (4.6) |
| Equestrian | 101 009 (3.2) | 82 475 (3.8) |
| Motor sports | 82 099 (2.6) | 61 524 (2.8) |
| Shooting | 79 986 (2.5) | 62 009 (2.8) |
| Cycling | 78 933 (2.5) | 61 110 (2.8) |
| Martial arts | 75 574 (2.4) | 46 957 (2.2) |
| (Other sports) | 983 369 (31.2) | 665 313 (30.5) |

Note: All memberships are all memberships lasting one day or longer. Active memberships are memberships lasting more than one season or with a paid licence. Age is measured as age at start of each membership. All other sports are included in S-Table 2.

Of all 3 148 213 memberships, 1 606 048 (51%) belonged to unique individuals, suggesting that, on average, a member had 2.0 memberships and 1.7 active memberships. Most memberships (69%) lasted more than 365 days. Among active memberships, 93% lasted more than 365 days.

Football was the most common sport, comprising 24% of all memberships. This was followed by handball (11%), gymnastics (9%), and skiing (7%). For active memberships, football remained the most common sport (25%), followed by handball (11%), with skiing (7%) being slightly more common than gymnastics (7%).

Table 1 shows descriptive characteristics of all and active memberships. In Table 1 age is measured as the age at the start of each membership. More than 40% of both all memberships and active memberships in NSMD started when the individual was 10–12 years. In Figure 1, in contrast, the unit of analysis is membership-year. This figure shows the age distribution for all individuals being members at least one day each year. The left panel shows the age distribution for all memberships and the right panel shows the age distribution for active memberships. The blue and red lines show smoothed distributions for males and females. Both all and active memberships had a similar distribution with a peak at age 11. Membership rates then rapidly declined until the age of approximately 30, with a slight increase for 40- and 50-year-olds.

**Figure 1.**
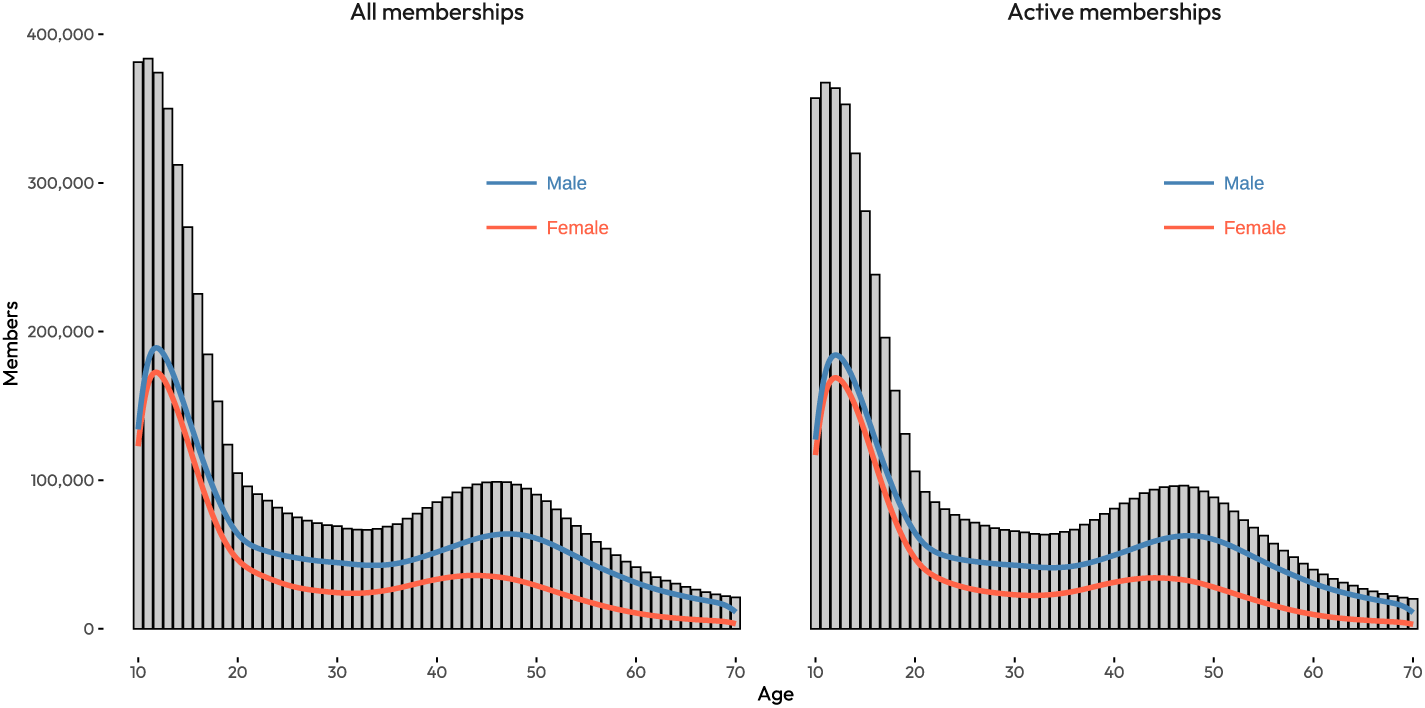
Age distribution of all and active memberships in NSMD. Note: Figure 1 shows all memberships (≥ 1 day) in the left panel and active memberships (≥ 1 season or paid licence) in the right panel. Each count represent a person-year being a member the year the member turns the given age.

### Coverage

The purpose of NSMD has since its establishment in 2015 been to cover all registered memberships in organized sports in Norway. Figure 2 shows the share of memberships found in NSMD compared to the total number of memberships reported by NIF. The upper panel (a) shows the coverage for all memberships, stratified by age groups. The lower panel (b) shows the coverage for active memberships. All six panels show that coverage has gradually increased from 2015 to 2023. It is also evident that coverage is highest for the youngest, 13–19 years. This is also the age group where sports participation is most common. The coverage is also higher for active memberships than for all memberships. For all membership the coverage reaches 85% in 2023 for 13–19 years, and 58-64% for the elder groups. For active membership the coverage approaches 93% for 13–19-year-olds in 2023, and 76% for the older age groups. In Figure 2, we included all sports, although golf was not thoroughly captured in the NSMD. After removing golf from both the numerator and the denominator, which was only possible for active memberships since NIF does not report all memberships stratified by sports federation, the coverage reached 96% in 2023 for 13-19-year-olds, 85% for 20-25-year-olds, and 89% for 26-70-year-olds (S-Figure 1). The total number of memberships in NIF’s reports and NSMD is shown in S-Figure 2.

**Figure 2.**
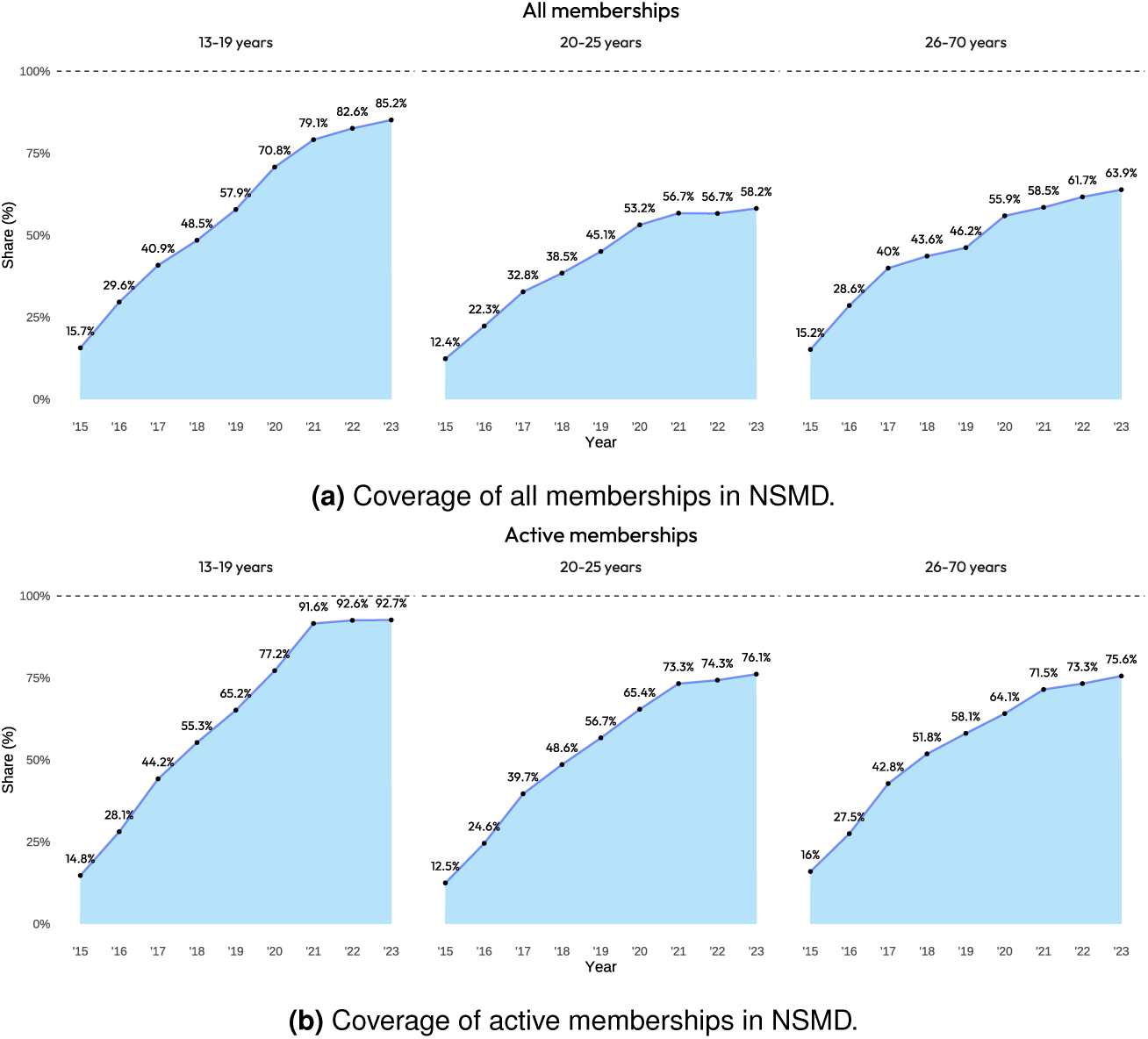
Coverage of NSMD.

Figure 3 shows the yearly coverage of the ten most common sports among 13–19-year-olds. While all ten sports had a coverage below 40% in 2015, all sports exceeded 80% coverage in 2023. For some sports, such as equestrian, gymnastics, and motorsport, the coverage exceeded 100% in 2023, illustrating that there were more registered members in NSMD than in NIF’s reports. Similar plots for all sports and all three age groups are provided in the Supplementary file (S-Figure 3).

**Figure 3.**
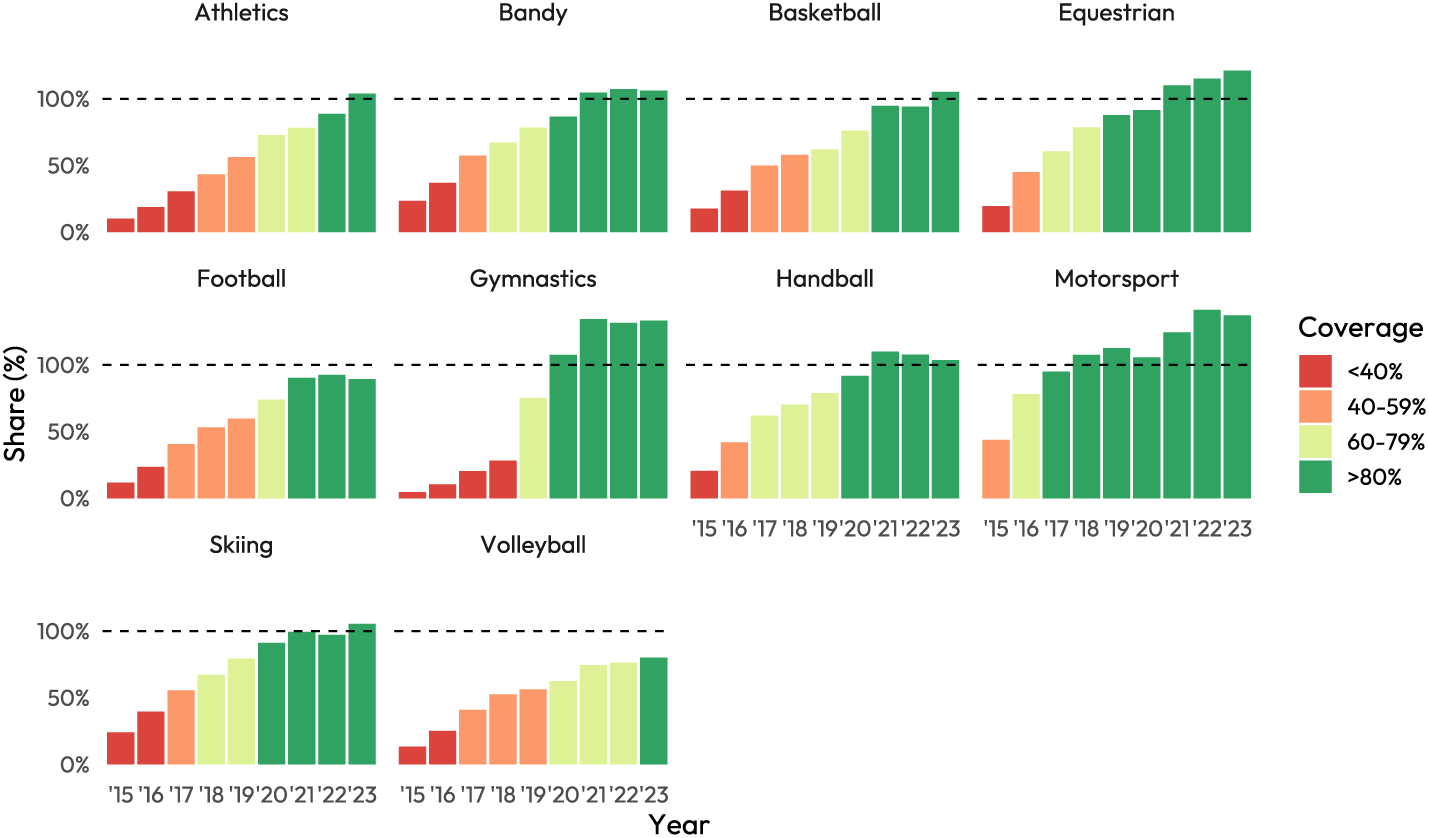
Coverage of top ten sports. Note: Coverage is estimated for active memberships among ages 13–19 between 2015 and 2023. Coverage is measured as the number of active memberships in NSMD divided by the number of active memberships from NIF’s reports. Coverage exceeding 100% suggests that there are more memberships in NSMD than in the reported numbers from NIF.

We observed only minor differences in member characteristics (age, geography, and parental income) in years with low coverage compared to years with high coverage (Supplementary Information 2).

## Discussion

Since its establishment in 2015, the NSMD has grown to include more than 3 million memberships and 1.6 million unique individuals, across 55 sports federations and 279 sports disciplines. In this paper, we have introduced two operational definitions of memberships for use in future research: (i) all memberships lasting at least one day, and (ii) active memberships, defined as participation over a full season or with a paid licence. Using the active definition, the dataset comprised more than 2 million active memberships.

The coverage of the database has improved substantially over time, increasing from approximately 10–15% of all memberships in 2015 to 60–90% of memberships in the years following 2021. Coverage was highest for 13–19-year-olds, and lower for 20–25-year-olds and 26–70-year-olds. This discrepancy was partly attributed to golf not being captured by NSMD. In reports by NIF, the top five sports in terms of active memberships were football, handball, golf, company sports, and skiing ^12^. However, golf and company sports were not fully captured in the NSMD. Only 7% of golfers were included due to incomplete integration of the golf federation’s membership system ^15^. Excluding golf greatly improved coverage for these groups from approximately 75% to 85-90% (S-Figure 1).

For certain sports and age groups, coverage exceeded 100%, indicating that more memberships were recorded in NSMD than reported in NIF’s annual statistics. When calculating coverage, we implicitly assumed that the numbers in NIF’s annual reports were correct. However, these figures were based on club submissions without a formal verification process. Clubs could find it challenging to count the precise number of members, and approximately 5% did not report membership figures to NIF at all ^12^. As a result, in later years, the available numbers in NSMD may offer a more precise depiction of the actual number of sports memberships in Norway than in NIF’s reports. However, determining which of these sources is the most reliable remains challenging.

The coverage of NSMD varies substantially over time. By 2021, coverages of most sports among 13–19-year-olds exceeded 90% (S-Figure 3), indicating the NSMD captured almost all memberships. Our analysis of demographic and socioeconomic distributions between years of low (<40%) and high (≥80%) coverage revealed only minimal discrepancies (S-Figures 4–6). These were primarily influenced by sport type rather than demographic or socioeconomic factors. This suggests that researchers may use, though with caution, the longitudinal data from e.g. 2015 to follow members over time without excluding particular socio-demographic groups.

A key strength of the NSMD is the inclusion of all organized sport memberships for the entire Norwegian population, which allows for population-wide analyses across a broad range of sports. The longitudinal nature of the data enables researchers to track exposures and outcomes over time, studying events before, during, or after joining organized sports. With the ability to link these data to other Norwegian registries and data sources through the unique national identity number of each individual, researchers can explore sports participation in relation to socio-economic, health-related, and other types of outcomes ^14^.

Our descriptive analysis of the NSMD showed that participation in organized sports was dominated by younger cohorts. Memberships peaked at age 11 for both boys and girls. This finding corresponded well with both NIF’s reports ^12^ and studies from other countries ^16^. As more than 90% of Norwegian children take part in organized sports at some point during their childhood ^17^, and organized sports constitute their largest source of physical activity ^18^, the NSMD is particularly valuable for studying children’s participation in sports. In contrast, participation patterns differ for older adolescents (*≥* 16 years) and adults. The largest dropout of organized sports occurs during lower secondary school and in the transition to upper secondary school ^19^, and surveys show that these age groups are more likely to engage in physical activity outside of organized sports, such as gyms ^20^. The NSMD data showed an increase in members for 40- and 50-year-olds, which may be a result of either a resurge in senior athletes, or the inclusion administrative members such as coaches, volunteers, or board members. The increase in members aged 40–50 years is also coherent with age distributions from NIF’s reports ^12^.

Although the strengths of the NSMD relate to the population-wide coverage, longitudinal aspects, and the ability to link it with other individual-level data, the data source also contains some limitations. The structure of the database leads to challenges such as overlapping information and duplicate entries, without the ability to determine whether these memberships, in fact, are overlapping or results of erroneous registrations. For example, an individual may be registered with one football membership lasting from January 1st, 2020 to January 1st, 2023, and another football membership from October 2nd, 2022 to January 1st, 2024. For most practical research, this could likely be considered one sustained membership. In addition, there may be uncertainty surrounding participation levels. The NSMD contains no information on physical activity, i.e., we do not know whether a member attends practices once every two weeks or seven days a week, and we do not know whether this differs between ages and sports. In the future, we plan to link the NSMD with large population based surveys, such as Trøndelag Health Study (HUNT) ^21^ or the Norwegian Mother, Father and Child Cohort Study (MoBa) ^22^, to validate whether sports memberships may be used as proxies for physical activity levels. Being a member does not necessarily reflect active participation, as individuals can stop attending practices or games before deregistering their memberships. We addressed this by incorporating licence data, which requires active payment to participate in games and tournaments, and we defined active memberships as those lasting at least one season or with a paid licence. Using all memberships and active memberships may serve to answer different questions. Studies of socio-economic barriers may include all memberships, whereas analyses of physical or psychosocial benefits of sustained participation may focus on active memberships.

A final limitation is that the period the NSMD reached high quality, in 2021, coincided with the covid-19 pandemic. During the pandemic, Norway imposed extensive restrictions on organized sports ^23,24^. Restrictions varied throughout the country, with stricter measures in urban areas with higher infection rates. Restrictions also differed between different sports, age groups, and levels of professional activity. Although NIF’s reports indicate a minor decrease in membership in 2020 and 2021^12^, NSMD saw an increase in memberships during the same period, likely due to increasing coverage (S-Figure 2). A survey in 2022 revealed that while participation in organized sports decreased during the pandemic, participation in the same sports increased after the pandemic ^17^.

Compared to previous research, the large majority of studies on organized sports participation are from surveys with self-reported data on participation. There are a few exceptions. For example, researchers from Australia have used de-identified registration data for a limited number of sports to investigate ages, genders, and whether sport members tend to live in urban or rural areas ^16^. Similarly, an ongoing initiative in Norway uses membership data with birth dates to investigate the relative age effect in sports ^25^. However, none of these projects involve the ability to link membership data to other registries on the individual level ^14^. Hence, NSMD represents the first possibility of these types of studies.

## Conclusion

The Norwegian Sports Membership Database (NSMD) contains all recorded and linkable memberships in organized sports in Norway from 2015 to 2024. From 2019, NSMD contains two-thirds of all active memberships, and around 90% between 2021 and 2023, for 13–19-year-olds. By linking NSMD to administrative health and sociodemographic registries, researchers can conduct longitudinal studies to investigate what happens before, during and after individuals participate in different sports.

## Contributions

FM, IHE, and KM designed the study. FM and IHE prepared the data. FM conducted all data analyses and drafted the manuscript. All authors (FM, IHE, JMK, AM, RKH, AF, KM) contributed to interpreting the results, and all authors revised the draft critically. Supervision was provided by KM. FM drafted the revision. All authors approved the final version of the manuscript to be published and agree to be accountable for all aspects of the work.

## Data Availability

The datasets generated and analyzed for the current study are not publicly available due to data protection reasons. Researchers wishing to use this data must contact the Norwegian Olympic and Paralympic Committee and Confederation of Sports. R-scripts are available on GitHub: https://github.com/MethiF/2025_The_Norwegian_Sports_Membership_Database

## Acknowledgements

We would like to thank Terje Kleiven, Ingvild Reitan, and Guro Aurtande at the Norwegian Olympic and Paralympic Committee and Confederation of Sports (NIF) for thorough help and assistance in using and understanding the NSMD data. We would also like to thank Kjetil Telle, Kristian Bandlien Kraft, and Runar Barstad Solberg at the Norwegian Institute of Public Health (NIPH) for valuable feedback and comments during the drafting of the manuscript.

## Supplemental material

This supplementary information contains the following:

### Supplementary information 1: Determining seasons in different sports

Before the 21st century, sports were often classified as either winter or summer activities. For example, the Norwegian athlete Bjørn Wirkola excelled in ski jumping during the winter while also playing football for Rosenborg in the summer. Similarly, Laila Schou Nilsen achieved remarkable success across several disciplines: she won an Olympic medal in alpine skiing, two world championships in speed skating, multiple Norwegian tennis championships, and several national titles in handball. While these achievements were nothing short of extraordinary, they would be considered almost impossible today. This is because most sports have now become year-round activities, with competitive seasons that rarely pause.

To define an active member as someone participating for the duration of a full season, it is necessary to clarify what is meant by a “season”. For most sports, there is no clear-cut definition of when seasons start or end. Practices, games, and tournaments often take place throughout the year. This was also our experience when talking with the different sport federations in Norway. However, in order to make a coherent measure of a season to check the coverage of the database, we based our definition on the timing of games, tournaments, and competitions. We categorized 52 sports (all 55 sports with the exception of company sports, multisports, and unversity sports) as having either a winter season (September–May) or a summer season (March–November). To ensure consistency, both summer and winter seasons were defined as spanning eight months, even though this may not perfectly reflect the competition period for every sport.

A member is considered a seasonal participant if:

- In winter sports: membership starts no later than the end of September in one year and continues beyond the beginning of May the following year.
- In summer sports: membership starts no later than the end of March and continues beyond the beginning of November the same year.

An exception was made for 2024, as membership data was censored on October 31st, 2024. In that case, ongoing memberships at the cut-off date were counted as active for summer sports. However, for winter sports, memberships that started before September 2024 but were only confirmed as ongoing on October 31st, 2024, were not defined as covering a full season.

When counting the number of active memberships each year, we added an additional restriction that each membership had to last a minimum of 90 days the given calendar year to ensure that sports with winter seasons were not counted twice.

For members aged 13 to 19 years in sports with available licence data, we ran a correlation analysis between being a member a full season and having a paid licence. This revealed that 52% of all season members had a paid licence, and that 74% of all licence holders were members for at least one season.

S-Table 1 lists all sports (federations), the disciplines within each federation, whether the database contains licence information, and the defined season (winter or summer).

**S-Table 1:**
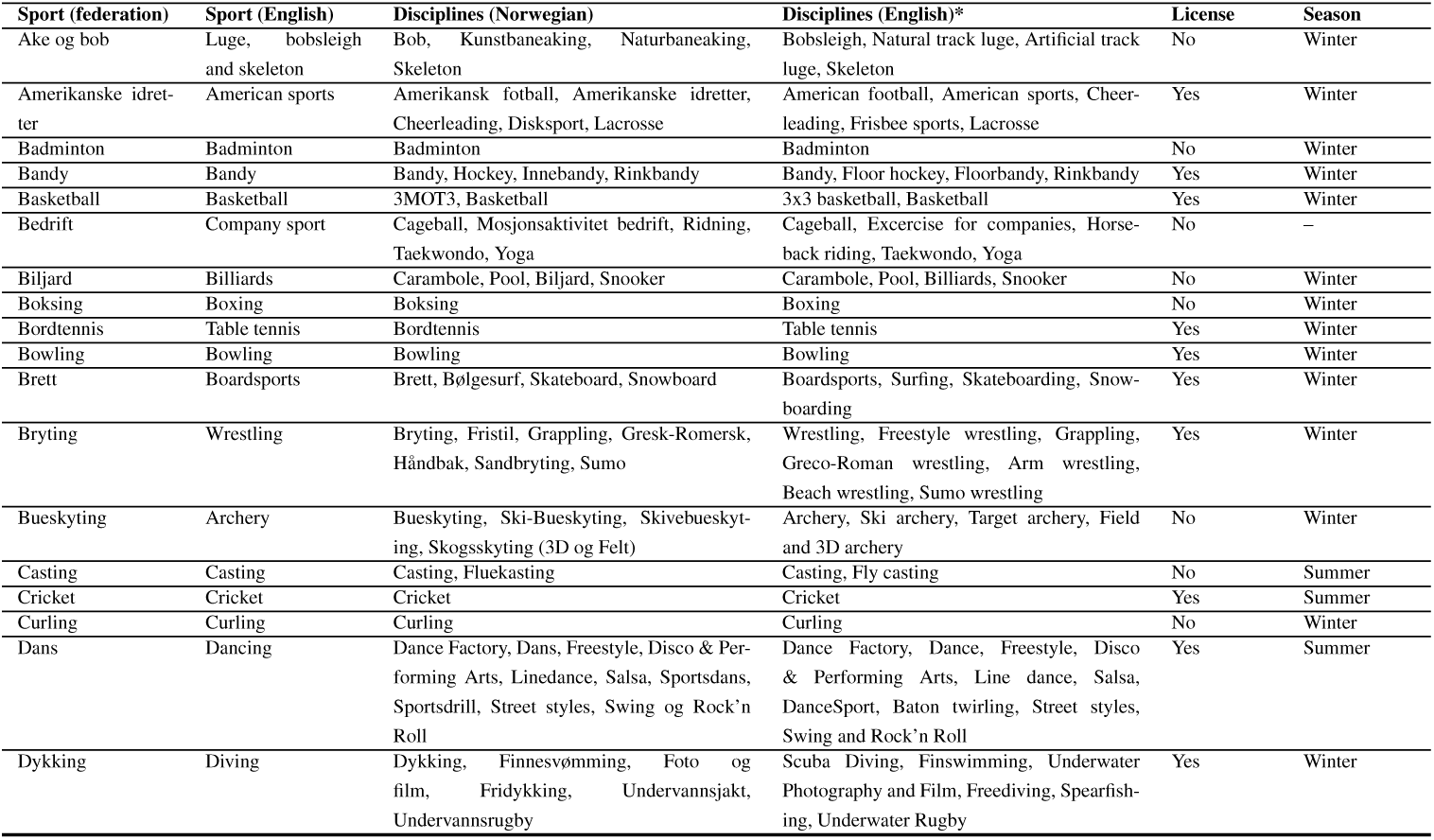
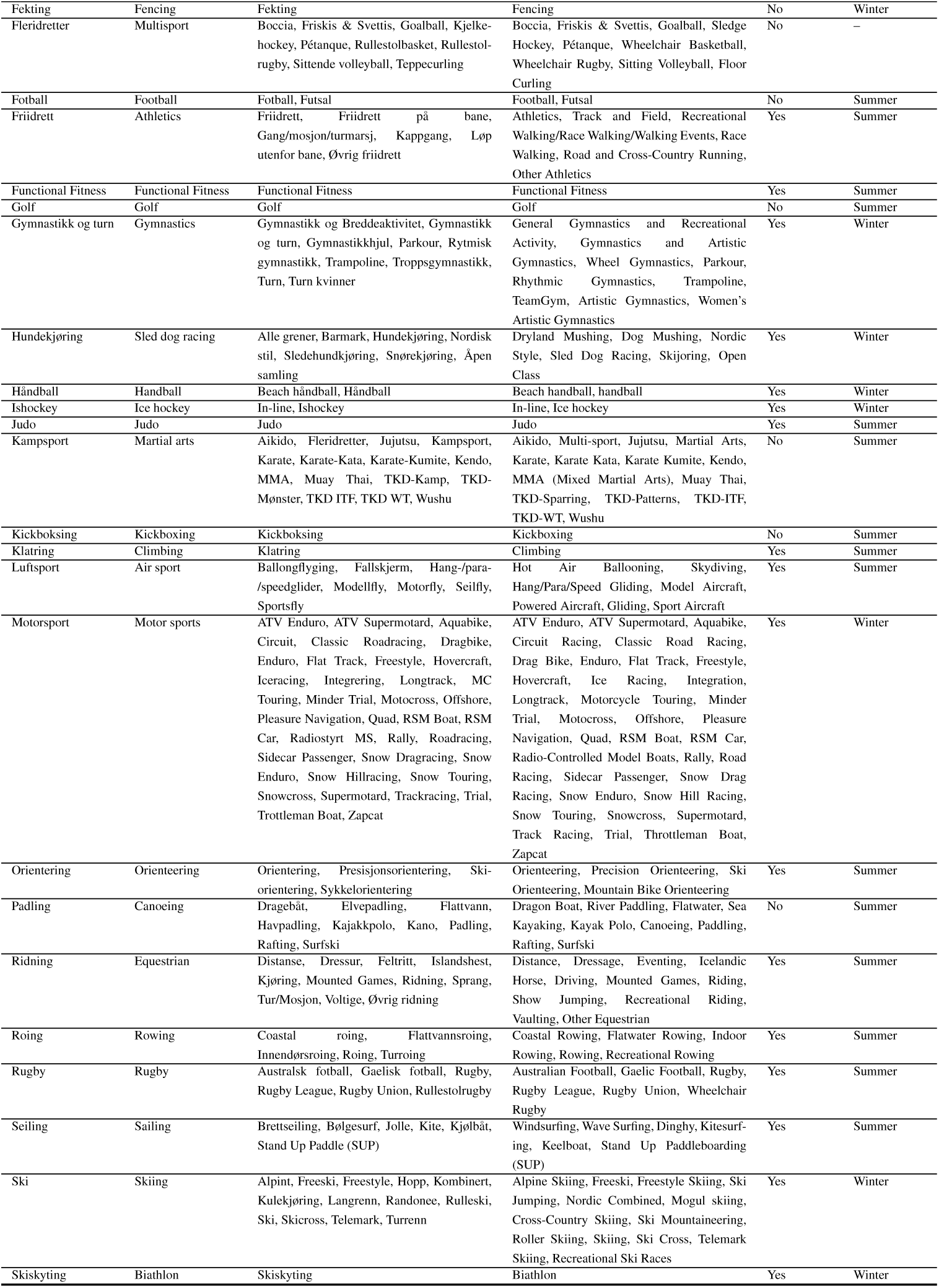
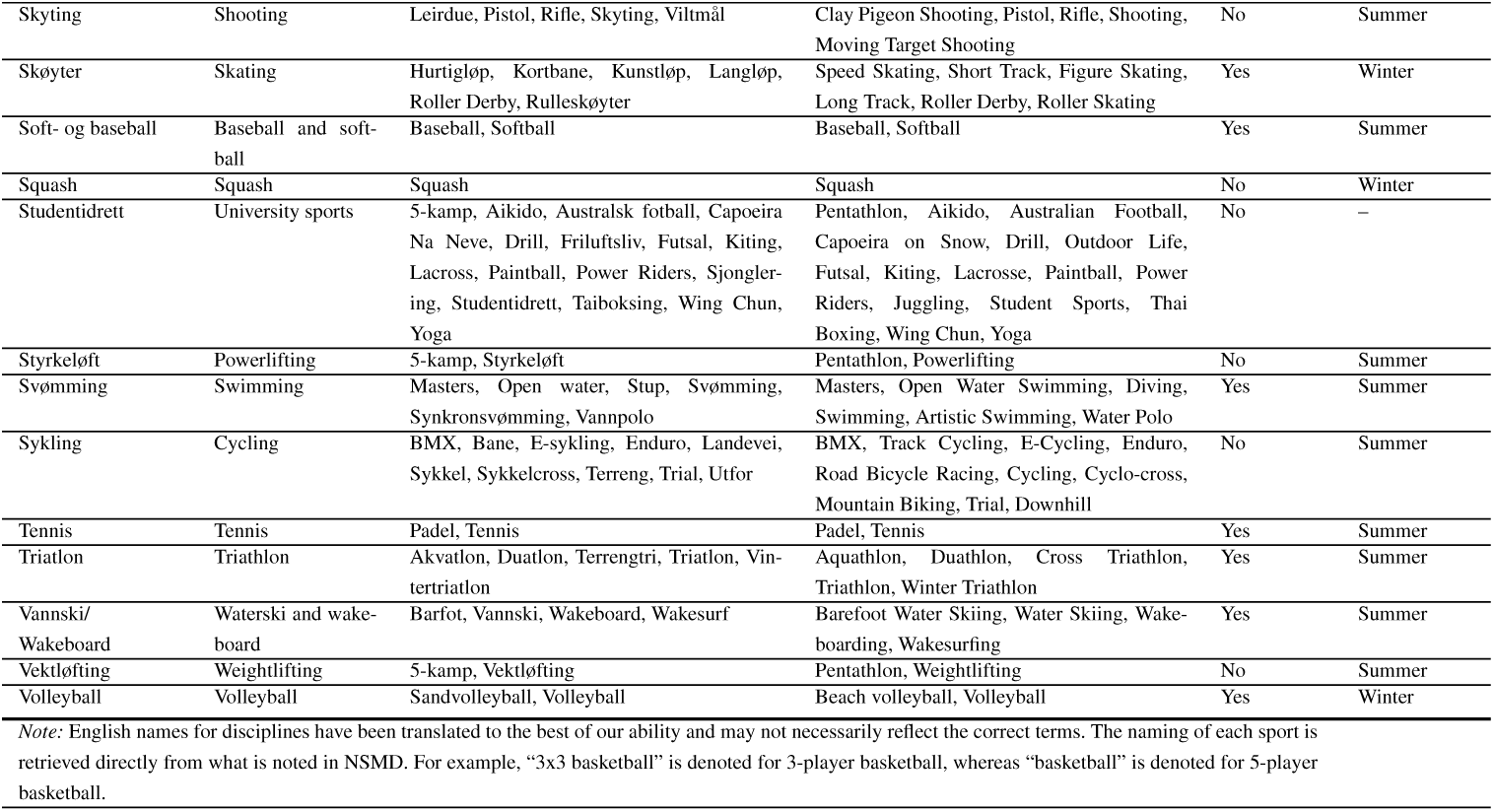
Sports, Disciplines, Licences and Seasons in NSMD.

### Omitted sports

When calculating coverage we removed company sports, student sports, and multisports. Company sports did not have reliable membership figures in either NIF’s reports or NSMD and were removed from both all and active memberships. Similarly, we removed university sports from the numerator and denominator in the analysis of active members as these members were included in sports associations (e.g., football in football) and not counted twice in NIF’s reported numbers, but they were only removed from the numerator in the analysis of all members, as they were not included in the denominator in NIF’s reports. Finally, we removed multisports from the analysis of active members as seasons were not definable (S-Table 1).

**S-Table 2:**
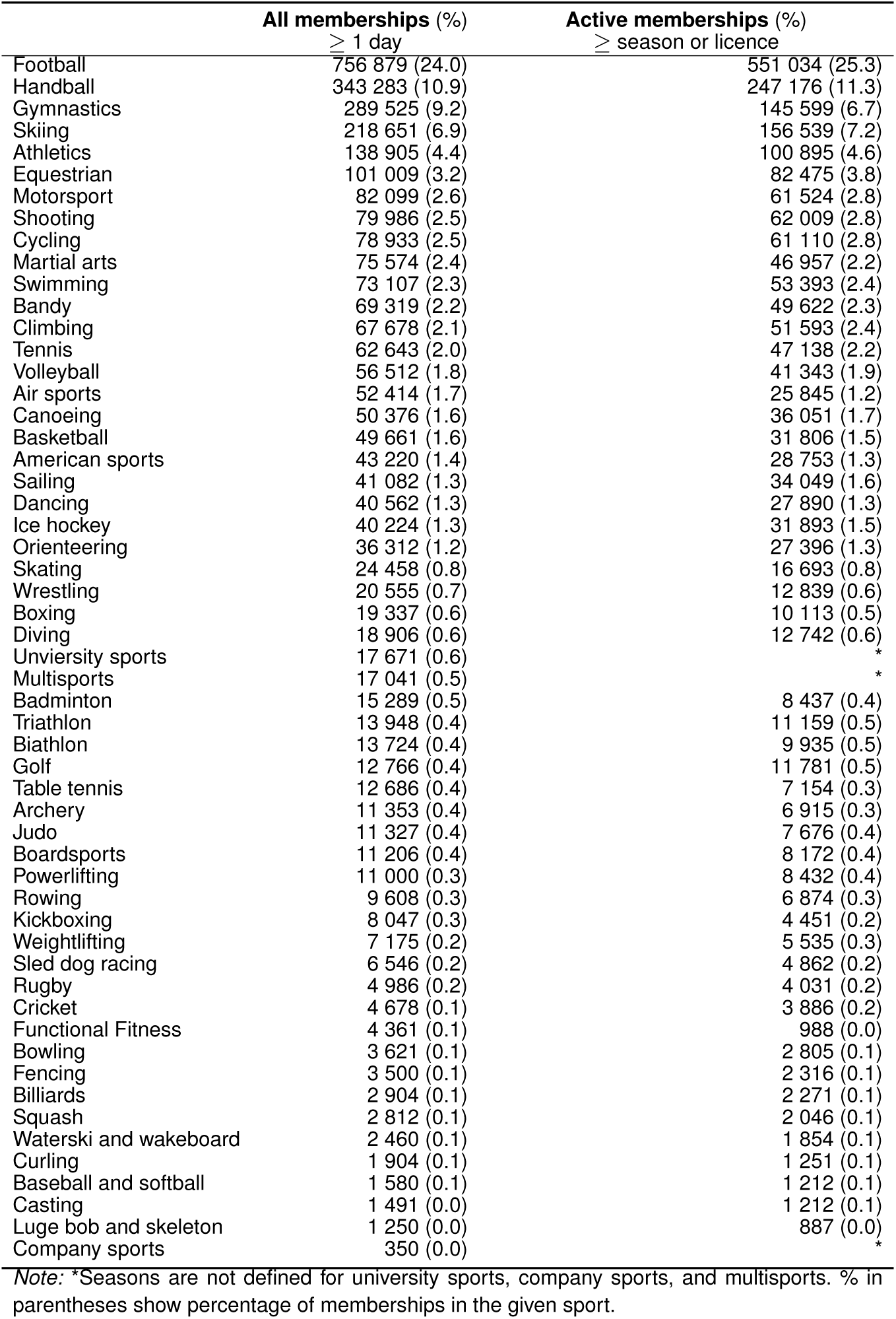
All and active memberships by sport.

### Supplementary information 2: Detailed on coverage

Using membership data from years with low coverage may produce biased estimates if systematic differences exist between members who are registered in low-coverage years and those in high-coverage years. An important question to consider is: if the coverage is only 20%, which groups are included in these 20%? For example, are these 20% only the oldest quintile, the wealthiest children, or participants from specific clubs or regions of the country? In such cases, estimates based on these years would be biased. However, if the missing memberships during these periods are missing completely at random (MCAR), the resulting estimates would remain unbiased.

To assess this, we compared the distributions of observable variables in years with high coverage (*≥* 80%) against years with low (<40%), fair (40–59%), and moderate coverage (60–79%). Specifically, we examined whether the type of sport, age, geographic distribution, and parental income of registered members remained consistent across different coverage levels, stratified by the ten most common sports. S-Figure 2 presents coverage for all 52 sport federations with defined seasons. The figure demonstrates that most sports reach coverage exceeding 80% in later years, but also reveals considerable variation between federations and ages. Golf, curling and function fitness did not exceed 60% coverage for any age group.

**S-Figure 1:**
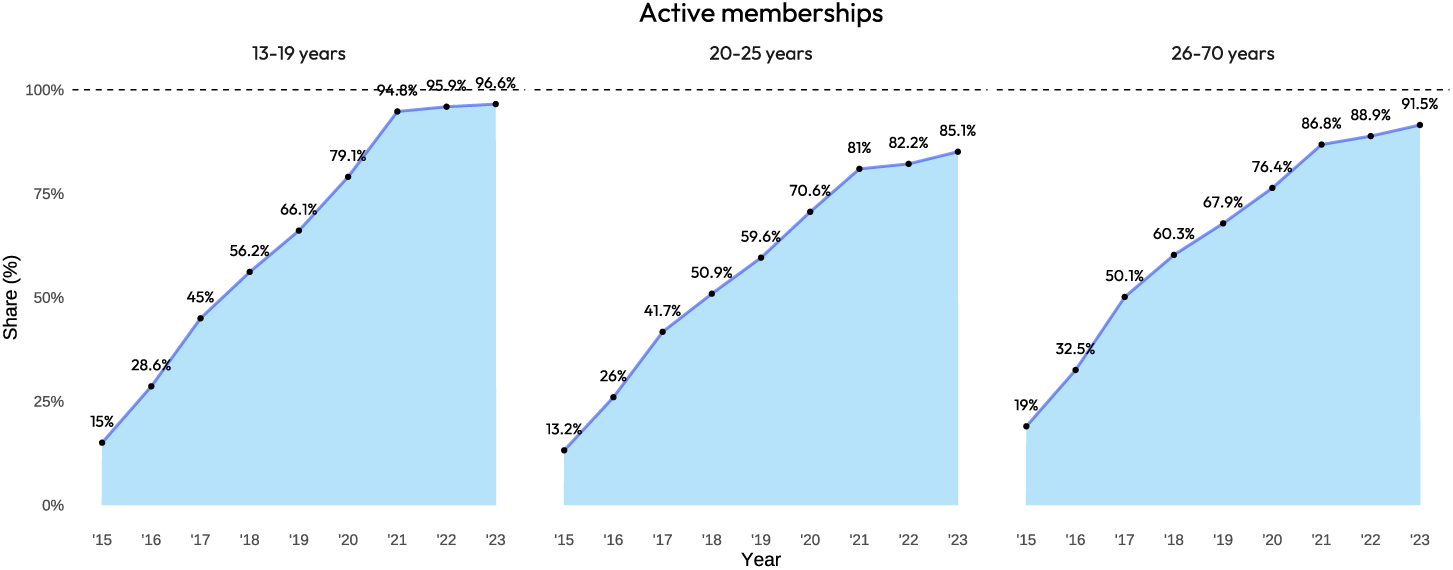
Coverage of active memberships in NSMD (without golf, company sports, university sports and multisports)

**S-Figure 2:**
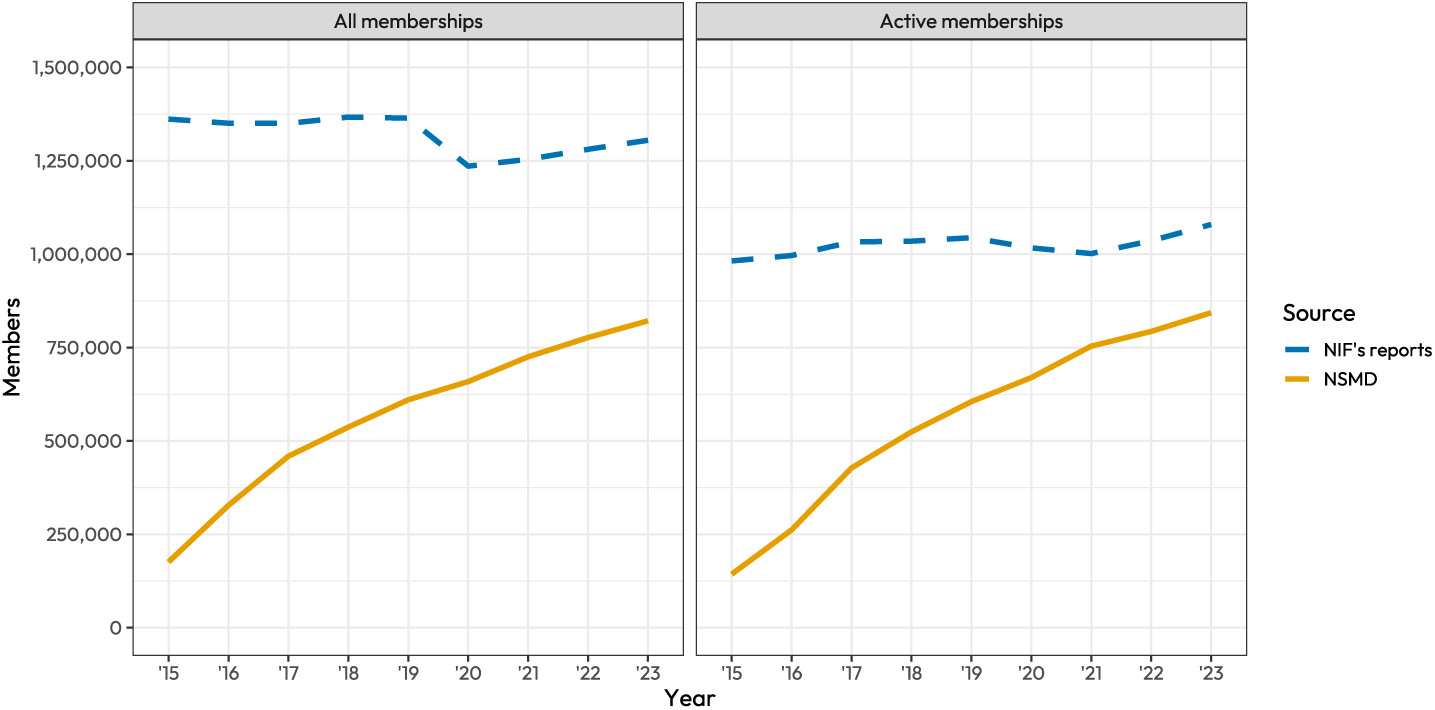
Total number of memberships in NSMD and NIF’s reports, 13–70 years.

For sociodemographic characteristics, we focused on the top ten sports among individuals aged 13–19. Visual inspections of distributions across years with varying coverage suggested that age, geographic location, and parental income remained relatively stable (S-Figures 3–5). Age distributions for the ten most common sports showed only minor variations (S-Figure 3).

**S-Figure 3:**
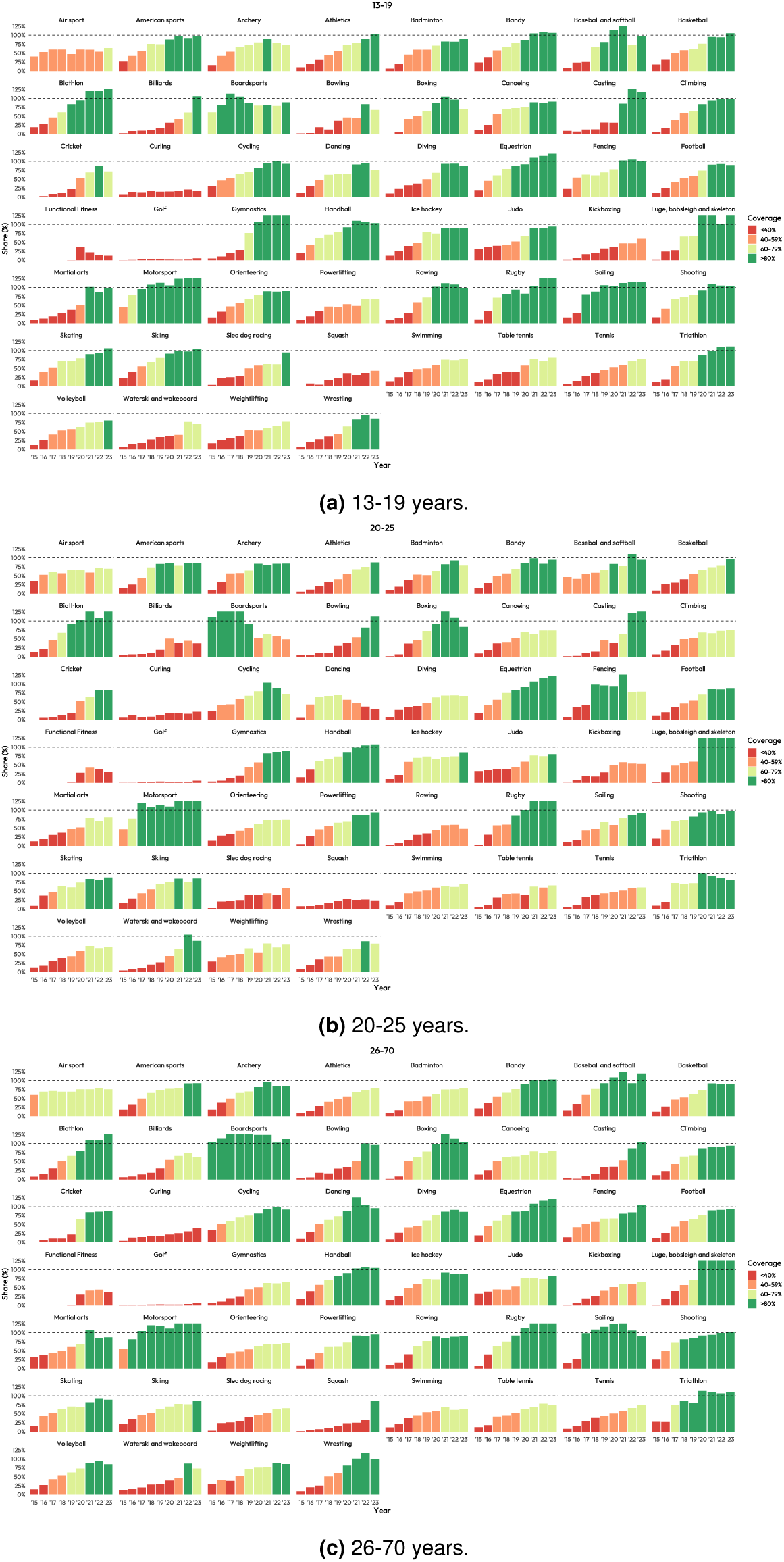
Coverage by type of sport and age groups.

**S-Figure 4:**
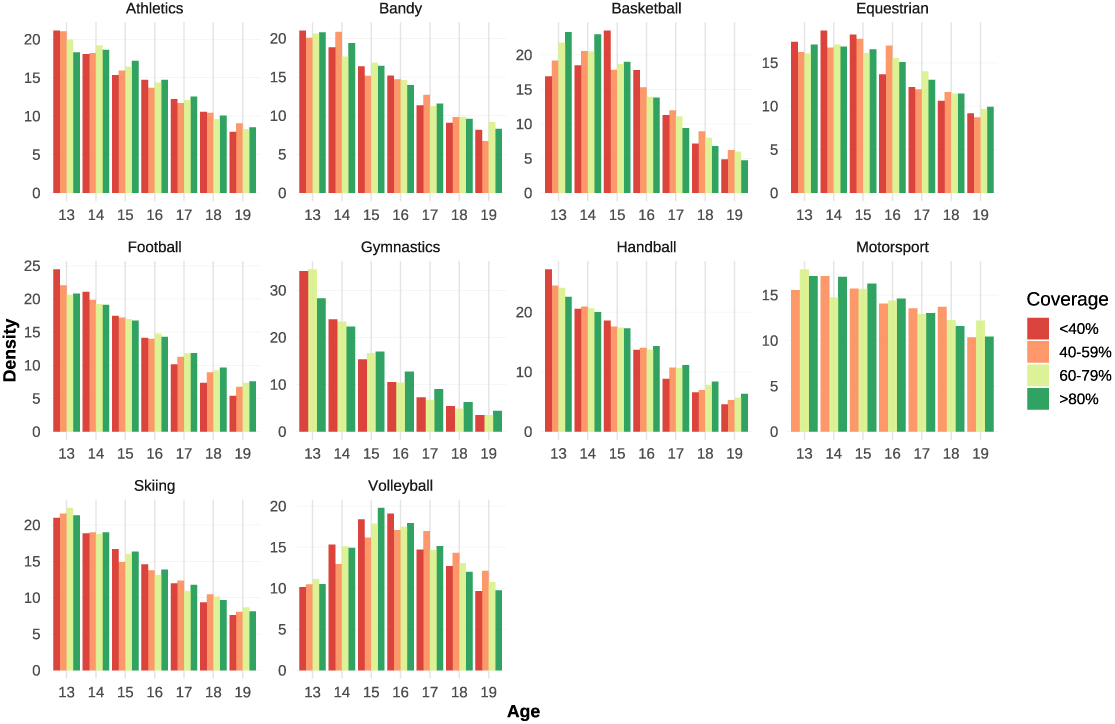
Coverage by age.

**S-Figure 5:**
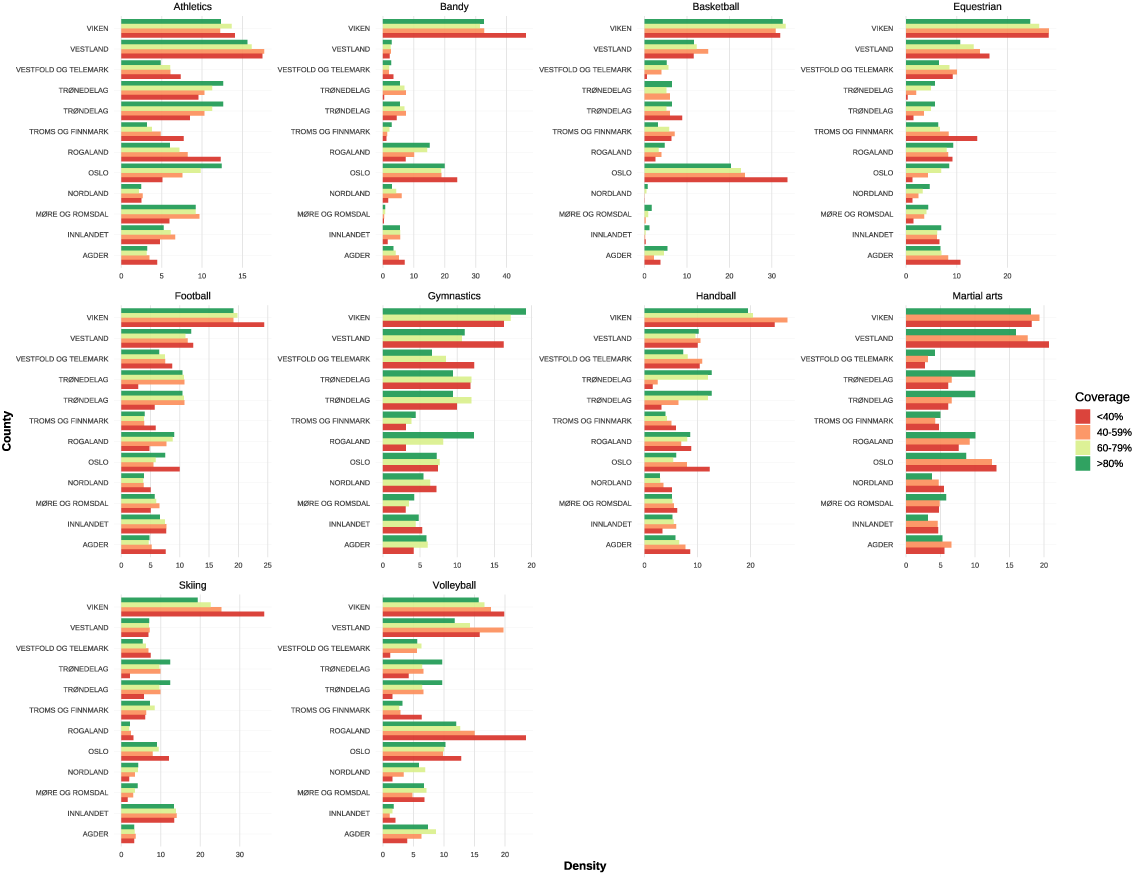
Coverage by geography.

**S-Figure 6:**
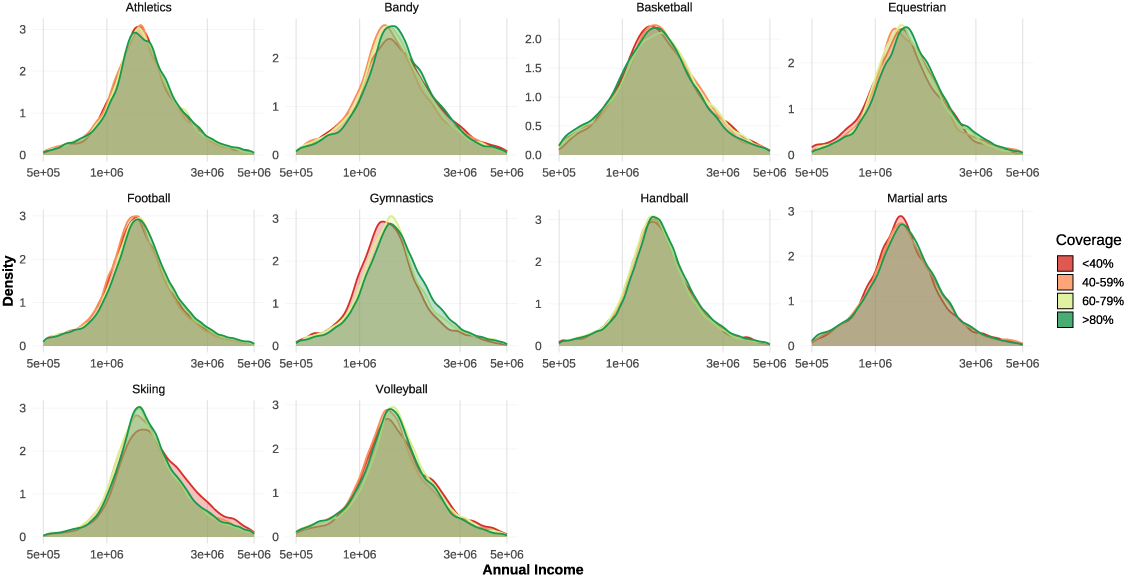
Coverage by income.

For geographic differences, we used Norwegian counties as the unit of observation. Between 2015 and 2024, Norway underwent several administrative reforms to its county structure. In this paper, we use the “2020 counties”, as these aligned more closely with the Norwegian Olympic and Paralympic Committee and Confederation of Sports (NIF)’s regional sports federations. All with the exception of Troms and Finnmark, which constitute a single county but are represented by separate regional sports federations. Within this framework, we observed that Viken was slightly overrepresented in skiing and bandy, Rogaland was slightly overrepresented in volleyball, and Oslo was slightly overrepresented in basketball during years with lower coverage quality (S-Figure 4). In contrast, Trøndelag was slightly underrepresented in skiing and handball, Oslo was slightly underrepresented in athletics, and Vestfold and Telemark, and Norland were slightly underrepresented in volleyball during years of low coverage. However, these patterns may also reflect genuine temporal shifts in participation trends. For example, in recent years there may have been relatively fewer skiers from Viken and athletes from Rogaland, and relatively more participants from Trøndelag and Oslo.

Parental income distributions appeared almost identical within sports across years with different coverage levels (S-Figure 5). Parental income was defined as the combined annual gross income of both mother and father, lagged by one year and adjusted for inflation to 2022 NOK. The stability of these distributions across years suggests that there are no major selection biases related to age, geography, or socio-economic status during periods of varying coverage quality.

We also conducted statistical t-tests that compared mean differences in age, geography, and income (not included). These tests revealed some significant disparities in specific sports. For example, in football and handball, mean age and income differed significantly across nearly all coverage-level comparisons, with the exception of football age when comparing years with good versus high coverage. Given that these are the two largest sports by membership, such results may partly reflect very small standard errors, which make even minor differences statistically significant. By contrast, in basketball (income) and skiing (age), mean differences were never statistically significant.

